# Alterations in Lipid Profiles in Children with Perinatally Acquired HIV Infection living in Ghana

**DOI:** 10.1101/2025.01.16.25320654

**Authors:** Ruth Ayanful-Torgby, Veronika Shabanova, Akosua A. Essuman, Emmanuel Boafo, Linda E. Amoah, Elijah Paintsil

**Affiliations:** Department of Pediatrics, Yale School of Medicine, New Haven, CT USA; Department of Immunology, Noguchi Memorial Institute for Medical Research, University of Ghana, Accra, Ghana; Department of Biostatistics, Yale School of Public Health, New Haven, CT USA; Department of Pediatrics, Boston University Chobanian & Avedisian School of Medicine, Boston, MA USA

**Keywords:** Dyslipidemia, children living with HIV, Lipids profile

## Abstract

Children and adolescents who acquired HIV perinatally and are on lifelong antiretroviral therapy (ART) are at increased risk of lipid abnormalities, premature atherosclerosis, and early onset cardiovascular diseases (CVD). Majority of these children reside in sub-Saharan Africa, where monitoring of lipid profiles is not routine. In this study, we assessed the age dependent prevalence of dyslipidemia among children and adolescents who acquired HIV in Ghana.

In this cross-sectional study we examined lipid profiles of 397 children aged 6 to 18 years. Dyslipidemia was defined by elevated total cholesterol (TC) (≥200 mg/dL) or triglyceride (TG) levels (>150 mg/dL) or low-density lipoprotein cholesterol (LDL-C) (>130 mg/dL) or reduced high-density lipoprotein cholesterol (HDL-C) (<35 mg/dL). Dyslipidemia prevalence, by at least one criterion and for each criterion separately, was compared between age groups 6 - 12 (pre-teen) and 13 - 18 (teenage) years, as well as by sex.

The overall prevalence of dyslipidemia was 42.32%, and by individual criterion was 9.07% using LDL-C, 11.84% using TC, 15.87% using TG, and 24.00% using HDL-C. Additionaly, 5.04% (n=20) of the participants showed abnormalities across three parameters. Teenagers had a higher overall prevalence of dyslipidemia (46.75% vs. 39.04%, p=0.12). Criterion-based dyslipidemia prevalence varied across age groups, with TC-based (14.04% vs. 8.88%, p=0.12) being higher in the pre-teen group, while elevated TG-based (20.12% vs. 12.72%, p=0.05) and low levels of HDL-based (28.99% vs. 20.60%, p=0.05) were more prevalent in the teenagers. The mean levels of TC (p=0.04) and LDL-C (p=0.03) were significantly higher in the pre-teen age groups. Females had nearly twice as high as that of males with prevalence of abnormal LDL-C levels (11.39% vs. 6.67%, p=0.13).

This study showed a high prevalence of dyslipidemia among children who acquired HIV perinatally and are on ART, with age-related variations in lipid profiles. These findings emphasize the importance of implementing routine monitoring for lipid disorders within this population.

## Introduction

Dyslipidemia, defined as increased low density lipoprotein (LDL-C), high triglycerides (TG) and cholesterol (TC), and low high density lipoprotein (HDL-C), is the most common metabolic disorder in individuals living with HIV, regardless of their treatment status ^1–3^. Moreover, there are reports of a significant association between dyslipidemia and the type of antiretroviral therapy (ART) regimens. For instance, non-nucleoside reverse transcriptase inhibitors (NNRTIs) and protease inhibitors (PIs) are associated with low HDL-C and hypertriglyceridemia ^4,5^. Before initiation of ART, HIV infection increases triglycerides and decreases TC, HDL-C, and LDL-C ^6^. There are mixed reports on the trends of the various lipids after initiating ART. In a study from Ethiopia, the prevalence of dyslipidemia was 70% and 58% for ART-experienced and ART-naïve children, respectively ^7^. In this study, the frequency of individuals with low HDL-C and hypertriglyceridemia were higher among children on ART, however, there were no differences in total cholesterol and LDL-C ^7^.

Dyslipidemia has been associated with a higher prevalence of cardiovascular diseases (CVD) among people living with HIV (PLWH) compared to the general population. The risk of CVD among PLWH is twice as high as those in the general population ^8,9^, primarily attributed to dyslipidemia. Moreover, CVD including conditions like myocardial infarction, manifests among PLWH a decade or two earlier than in the general population ^10^. This trend is particularly significant in sub-Saharan Africa, where HIV prevalence is high ^11^. There is a paucity of studies on CVD in children living with HIV, although they are susceptible to cardiovascular complications due to dyslipidemia ^12^. In a study of Ugandan children and adolescents living with HIV aged 1 to 17 years, the prevalence of dyslipidemia was 74%; of those with dyslipidemia, 56% had low HDL-C, 22% had hypertriglyceridemia, 16% had high LDL-C, and 11% had high total cholesterol levels ^190^. Elevated LDL-C and low HDL-C in children and young adults have been attributed to abnormalities in the arterial aorta ^13^, which serve as precursor to atherosclerosis ^14^. Additionally, low HDL-C levels are associated with inflammation, contributing to insulin resistance and other cardiometabolic disorders, including metabolic syndrome. Despite the established link between dyslipidemia and CVD, lipid profiles are not routinely monitored in children living with HIV, potentially hindering early detection and intervention effort. Considering that alteration in lipid metabolism is associated with cardiometabolic diseases, an emerging public health problem in most developing countries, including Ghana, it is imperative to detect precursors of these diseases, such as dyslipidemia. Early detection of dyslipidemia can help avert related pathologies, such as myocardial infarction and type 2 diabetes, which are on the rise in young adults living with HIV.

Taken together, dyslipidemia in children living with HIV is an emerging public health concern as continued lipid abnrmalities may contribute to the development of atherosclerosis and other CVD in early adulthood. Given the significant HIV burden in sub-Saharan Africa, a large proportion of children living with the virus in this region may be at risk of developing CVD associated with dyslipidemia, which could go unnoticed due to insufficient screening. To address this gap, our study aimed to assess the prevalence of dyslipidemia and its association with age and sex among perinatally HIV-infected children receiving ART in Ghana.

## Methods

### Study Participants

This multi-center cross-sectional study was conducted from 5^th^ January to 28^th^ June 2022 among children and adolescents living with perinatally acquired HIV, aged 6 to 18 years from study participants were recruited from ten pediatric antiretroviral treatment clinics in ten facilities in Greater Accra and Eastern regions of Ghana. Details of the study participants have been published previously ^15^. In brief, the study recruited 400 participants from ten pediatric ART treatment clinics across ten health facilities in Greater Accra and Eastern Regions of Ghana, which share similar socioeconomic indices. All participants acquired HIV pernatally and had been on uninterrupted antiretroviral therapy for at least a year, adhering to the national ART treatment guidelines for children and adolescents. These guidelines prioritize first-line, second-line, or third-line regimens with appropriate dosages based on ART class and age. We excluded all individuals with viral hepatitis and TB due to potential interference with lipid/glucose metabolism caused by alternative medications and regimens ^16^. All individuals who were clinically stable and willing to participate were recruited after being briefed on the study’s objectives and procedures. Age-appropriate written parental informed consent, assent, and consent were obtained prior to participants’ enrollment, followed by scheduled sampling procedure (blood draw and urine collection), questionnaire administration and anthropometric measurements taken at the clinics.

### Ethical Approvals

The study protocol was reviewed and approved by the Institutional Review Boards of the Ghana Health Service (Protocol # GHS-ERC: 025/09/21) and the Noguchi Memorial Institute for Medical Research, University of Ghana (Protocol # CPN 015/21-22). Written parental informed consent was obtained for children aged 6–12 years, while both parental consent and assent were obtained for participants aged 12–17 years. For participants aged 18 years, individual consent was obtained. All participants were assured of anonymity and data protection. Research data was stored and used in compliance with the ethical guidelines approved for the study, ensuring participant protection as previously reported ^15^.

### Data Collection

Detailed information on data collection methods has been published previously ^15^. In brief, we administered questionnaires to participants or their parents/guardians to collect demographic (age, sex, and residental location), educational (participants’ education), and socioeconomic information (parent or guadran educational level and occupation). Additionally, anthropometric measurements (upper arm circumference, waist circumference, hip circumference, and neck circumference) were conducted by a trained member of the study team or health staff.

### Biochemical Parameters

All participants provided 2.5 ml blood samples after an overnight fasting period, before the next morning’s blood collection. Serum was separated from 2 mL whole blood samples collected in gel tubes for subsequent analysis of lipid profile and the remaining 0.5 mL blood used for other laboratory investigations. The levels of TG, HDL-C, and total TC were measured using a chemical analyzer (Mindray BS-240, China). LDL-C levels were determined by the Friedewald formula ^17^.

### Definitions of Variables

The primary outcome of interest was dyslipidemia, defined as presence of any of the following parameters: by elevated TC (≥200 mg/dL), TG (≥150 mg/dL), or LDL-C (≥130 mg/dL), or reduced levels of HDL-C (≤35 mg/dL) ^18^. Age was stratified into two categorized as 6 - 12 years (pre-teen) and 13 - 18 years (teenage), while sex was categorized as female or male, based on gender assigned at birth.

### Statistical Analysis

Categorical variables such as sex, educational level, and caregiver type are summarized as count (n) and percentage (%), while numeric variables such as age were expressed as means and standard deviations (SD). Participant characteristics were compared between the two age groups using Student’s t-test or Chi-square statistics for continuous and categorical variables, respectively. We estimated the prevalence of dyslipidemia, defined as having at least one abnormal parameter, as well separately based on each of the four lipid parameters (TC, TG, LDL-C, HDL-C), and compared them based on age group (pre-teen vs. teenage) and sex (male vs. female) using two-samples z-test, summarizing as counts and percentage. Since we did not power our statistical analysis *a priori*, we did not rely solely on the strict two-sided alpha level of 0.05 to make conclusions, but coupled our interpretations with the magnitude of the observed effect sizes. For example, a difference of at least 5% in the prevalence of dyslipidimia or a relative difference of at least 1.5 (e.g., the prevalence in one group divided by the prevalence in a comparison group), was considered clinically meaningful. All statistical analyses were performed using R (version 4.2.2) and GraphPad Prism (version 9.01).

## Results

### Characteristics of Study Participants

The analysis included 397 of 400 participants with complete lipid profile data on four key lipid parameters, i.e., TC, TG, LDL-C, and HDL-C. Participants in the pre-teen age group (6 - 12 years) represented 57.43% of the study participants. Distribution of sex variable was similar in the two age groups. Variations in BMI measures were evident across the age groups (p<0.0001), with the pre-teen group with a mean BMI of 15.65 (SD=3.07) compared to 17.58 (SD=3.48) among the teenage group. The mean BMI Z-score values also differed (p<0.0001), with the pre-teen group showing a mean negative score of −0.48 (SD=0.66), while the teenage group demonstrated a mean positive score of 0.65 (SD=1.02) (**Table 1**).

**Table 1.** Characteristics of Study Participants.

| Variable | Total (N=397) | Age groups |  |
| --- | --- | --- | --- |
|  |  | Pre-teen (n=228) | Teenage (n=169) |
| Age, year, Mean (SD) | 11.57 (3.49) | 9.66 (2.51) | 16.94 (3.85) |
| BMI Mean (SD) | 16.94 (3.85) | 15.65 (3.07) | 17.58 (3.48) |
| BMI Z-score Mean (SD) | 0.00 (0.71) | -0.48 (0.66) | 0.65 (1.02) |
| WHR Mean (SD) | 0.83 (0.08) | 0.86 (0.07) | 0.80 (0.07) |
| **Sex, n (%)** |  |  |  |
| Female | 202 (50.88) | 117 (51.32) | 85 (50.30) |
| **Education, n (%)** |  |  |  |
| Basic | 354 (89.17) | 227 (99.56) | 127 (75.15) |
| Secondary | 41 (10.33) | 1 (0.44) | 40 (23.67) |
| None | 2 (0.50) | 0 (0.00) | 2 (1.18) |
| **Guardian, n (%)** |  |  |  |
| Parent | 305 (76.83) | 181 (79.39) | 124 (73.37) |
| Sibling | 12 (3.02) | 3 (1.32) | 9 (5.34) |
| Grandparent | 31 (7.81) | 15 (6.58) | 16 (11.24) |
| Other relative | 25 (6.30) | 13 (5.70) | 12 (7.10) |
| <sup>a</sup>Others | 24 (6.05) | 16 (7.02) | 8 (4.73) |

<sup>a</sup>Self or non-relative including living in orphanage Abbreviations: SD=standard deviation, BMI=Body Mass Index, WHR=Waist-Height Ratio

### Prevalence of Dyslipidemia among the Study Participants

One hundred and sixty eight participants had dyslipidemia, representing an overall prevalence of 42.32% in the study participants. Categorization of dyslipidemia based on the frequency of altered levels among the four lipid parameters showed that the majority of participants (n=114, 28.72%) had altered levels in only one lipid parameter. Thirty-four individuals (8.56%) showed abnormalities in two parameters, while a smaller subset of 20 individuals (5.04%) had abnormalities in three parameters. No individual showed dyslipidemia characterized by abnormalities in all four lipid parameters. Compared to pre-teens, teenagers had higher prevalence of at least one altered parameter of dyslipidemia (46.75% vs. 39.04%, p=0.12; **Table 2**). The prevalence of dyslipidemia and the number of altered lipid parameters did not differ between males and females, with the exception of proportion of females with abnormal LDL-C levels being nearly twice as high as that of males (11.39% vs. 6.67%, p=0.13; Supplementary **Tables S1 and S2**).

**Table 2.**
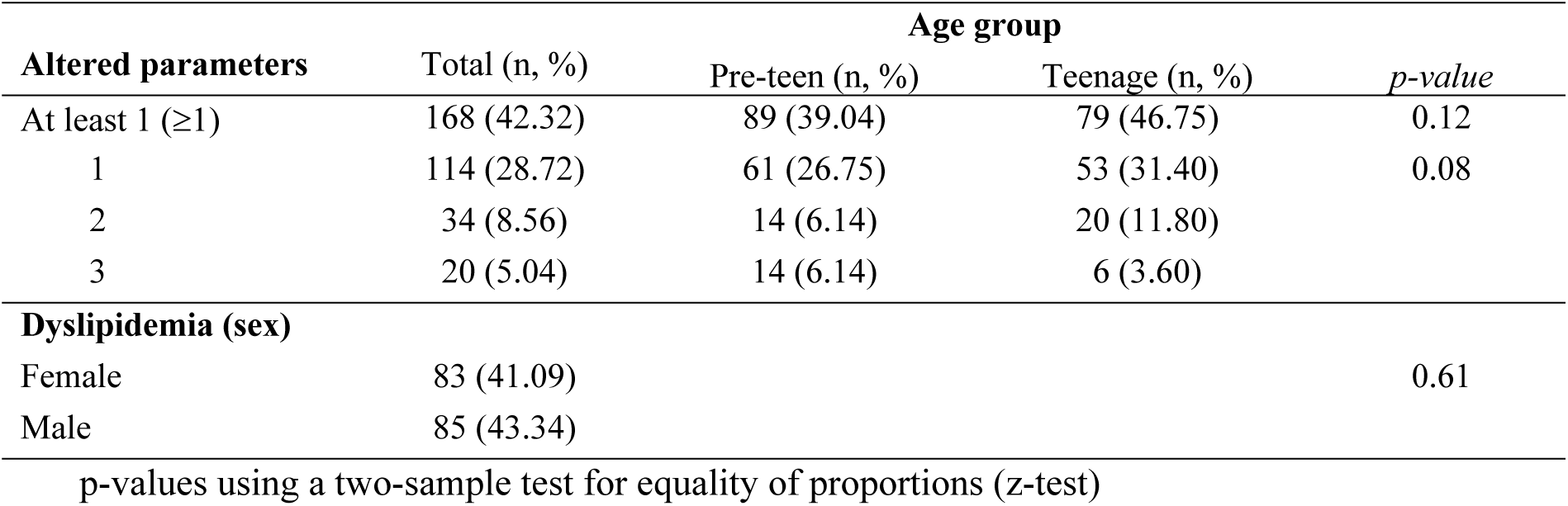
Comparison Frequency of Altered Lipid Parameters between Age Groups/Sex.

| Altered parameters | Total (n, %) | Age group |  | *p-value* |
| --- | --- | --- | --- | --- |
|  |  | Pre-teen (n, %) | Teenage (n, %) |  |
| At least 1 ($\geq 1$) | 168 (42.32) | 89 (39.04) | 79 (46.75) | 0.12 |
| 1 | 114 (28.72) | 61 (26.75) | 53 (31.40) | 0.08 |
| 2 | 34 (8.56) | 14 (6.14) | 20 (11.80) |  |
| 3 | 20 (5.04) | 14 (6.14) | 6 (3.60) |  |
| **Dyslipidemia (sex)** |  |  |  |  |
| Female | 83 (41.09) |  |  | 0.61 |
| Male | 85 (43.34) |  |  |  |

p-values using a two-sample test for equality of proportions (z-test)

### Age-Related Disparities in Lipid Profiles

When comparing lipid profiles across age groups (**Table 3**), the teenage group showed higher mean TG levels compared to the pre-teen group (p=0.15). LDL-C (p=0.03) and TC (p=0.04) levels were lower in the older (teen) age group compared to the younger population. However, the mean HDL-C levels between the two age brackets were similar (p=0.28). The prevalence of elevated TG was higher in the teenage group (p=0.05). The prevalence of elevated TC was nearly double in the younger age group compared to the older age group (p=0.12). Dyslipidemia prevalence based on LDL-C was similar in the two age groups (p=0.41). Additionally, the number of individuals with low levels of HDL-C was higher in the teenagers than in the pre-teen group (p=0.05).

**Table 3.** Lipid Profile and Comparison Across Age Groups.

| Parameter | Total (N=397) | Age Group |  | *p-value* |
| --- | --- | --- | --- | --- |
|  |  | Pre-teen (n=228) | Teenage (n=169) |  |
| **Mean levels (SD)<sup>b</sup>** |  |  |  |  |
| TC (mg/dL) | 156.07 (45.19) | 160.11 (47.09) | 150.61 (42.02) | 0.04 |
| TG (mg/dL) | 96.88 (56.17) | 93.37 (53.53) | 101.62 (59.50) | 0.15 |
| LDL-C (mg/dL) | 66.98 (52.15) | 71.87 (52.19) | 60.39 (51.52) | 0.03 |
| HDL-C (mg/dL) | 45.05 (16.00) | 45.81 (15.26) | 44.03 (16.93) | 0.28 |
| **Dyslipidemia, n (%)<sup>c</sup>** |  |  |  |  |
| TC (mg/dL) | 47 (11.84) | 32 (14.04) | 15 (8.88) | 0.12 |
| TG (mg/dL) | 63 (15.87) | 29 (12.72) | 34 (20.12) | 0.05 |
| LDL-C (mg/dL) | 36 (9.07) | 23 (10.09) | 13 (7.69) | 0.41 |
| HDL-C (mg/dL) | 96 (24.18) | 47 (20.61) | 49 (28.99) | 0.05 |

<sup>b</sup>p-values using Student's t-test, <sup>c</sup> p-values using two-sample z-test. Abbreviation: low density lipoprotein (LDL-C), high triglycerides (TG), cholesterol (TC), low high density lipoprotein (HDL-C)

## Discussion

A concerning increasing trend in the cluster of metabolic disorder components, including dyslipidemia, has been noted among children living with HIV, predisposing them to an increased risk to cardiovascular disease ^19^. In this study, we examined alterations in four lipid profile parameters: elevated levels of TC, TG, LDL-C, and low levels of HDL-C, all of which are acknowledged as significant contributors to CVD risk. Our findings revealed an overall dyslipidemia prevalence of 42.32% within our study participants, with the prevalences of individual parameters being 9.07% for LDL-C, 11.84% for TC, 15.87% for TG, and 24.18% for HDL-C. We observed that compared to the preteens in our study, teenage participants had higher prevalence of dyslipidemia primarily driven by the elevation in TG and depression in HDL-C. Dyslipidemia among the pre-teens was characterisized by elevated TC. The prevalence of dyslipidemia or levels of any of the four lipid parameters were not affected by being biologically male or female.

The prevalence of dyslipidemia in our study was high, however, a study conducted among South African children and adolescents living with HIV reported an even higher prevalence of 74% ^20^. In our study cohort, alterations in specific lipid parameters included 9% for elevated LDL-C, 12% for elevated total cholesterol (TC), 16% for elevated triglycerides (TG), and 24% for low HDL-C; this is consistent with trends observed in other African studies ^21–23^. Among the four lipid parameters measured, the prevalence of low HDL-C levels was the highest, followed by hypertriglyceridemia. It is well established that low HDL-C and hypertriglyceridemia are the predominant components of dyslipidemia in children living with HIV, with prevalence rates in sub-Saharan Africa ranging from 0% to 29% for hypertriglyceridemia ^24,25^, and 5% to 72% for low HDL-C ^22,25,26^. Similar findings have been reported in studies of adults living with HIV in Africa ^27^, although the prevalence in children tends to be lower, ranging from 6% to 9% for hypertriglyceridemia and 6% to 47% for low HDL-C^21–23,28^. Notably, a recent study in children living with HIV in Ethiopian reported a much higher prevalence of 72% for low HDL-C ^26^. Outside of Africa, higher rates of low HDL-C have also been observed among individuals living with HIV in Asian populations. For example, a recent study found that 47% of Thai youth aged 15 to 24 years had low HDL-C levels, while a study from India reported that 83% of ART naïve children living with HIV aged 2 to 12 years had low HDL-C ^29,30^. In our study, sex did not significantly influence lipid levels or the prevalence of dyslipidemia, except for LDL-C, which was more prevalent in females. Our finding is consistent with previous findings that females usually have higher LDL-C levels than males at puberty^31^. Our results are consistent with previous research examining dyslipidemia by sex in children living with HIV^32^. However, in the general population with respect to age or sex, lipid profiles have varied across studies and populations, particularly when factors such as obesity is considered ^33,34^.

We observed differences in TG, LDL-C, and TC levels between different age groups. Dyslipidemia prevalence varied across age groups, with most individuals showing abnormalities in only one parameter. The younger age group showed much higher levels of the lipid parameters (TC and LDL-C) than the older group as previously reported eventhough there are conflicting reports based on the population being studied ^35,36^. This difference could be attributed to confounding factors such as nutrition, viral load, or immune parameters, which were not measured in this study. While the prevalence of dyslipidemia was high across both age groups, teenagers were more likely to have altered levels of TG which is consistent with previous studies suggesting an increase in the prevalence of dyslipidemia during adolescence, possibly attributed to hormonal changes, or prolonged exposure to HIV and its treatments ^3,37^.

In terms of the frequency of dyslipidemia based on number of alterations in the four lipid parameters, we observed that the majority of participants showed abnormalities in just one lipid parameter as reported in other studies. However, close to 14% of our study participants showed alterations in two or three parameters concurrently. Understanding this variability and the extent of multiple markers of dyslipidemia is crucial for tailored interventional strategies, as different lipid abnormalities may confer varying cardiovascular risks. The presence of multiple-markers of dyslipidemia may not only increase the risk but also predispose individuals to coronary heart disease ^38,39^. Several studies have reported that disruptions in multiple lipid parameters significantly elevate CVD risk, with others citing the presence of low HDL-C levels as a key determinant ^41,42^. Low HDL-C level is a crucial determinant of cardiovascular risk because HDL-C plays a pivotal role in cholesterol metabolism, inflammation regulation, endothelial function, and the prevention of atherosclerosis. Therefore, the prevalence of dyslipidemia, particualarly low HDL-C, warrants routine monitoring of lipid profile in children living with HIV curb CVD sequelae in their adulthood.

Although our study provided valuable insights on dyslipidemia in children living with HIV in Ghana, there were limitations that need to be acknowledged. First, missing variables of HIV diseases status such as treatment history, ART regimen, viral load, and CD4 count. Second, the cross-sectional design and lack of HIV-specific factors could introduce bias and influence dyslipidemia prevalence estimates and other risk factors, as previously acknowledged ^15^. Third, the study design limits the ability to draw causal inferences. Furture longitudinal studies are needed to validate our findings.

In conclusion, our results show a high prevalence of dyslipidemia among perinatally children living with in Ghana. Low HDL-C levels, a pivotal predictor of CVD risk and inflammation, was the prevalent parameter among individuals with dyslipidemia in our study. This highlights the need for routine monitoring of lipid profile of children living with HIV and proactive management to prevent CVDs in adulthood. Furthermore, there is urgent need for further research into the etiopathogenesis of rising dyslipidemia and evolving epidemic of cardiometabolic diseases in children living with HIV, particularly in sub-Saharan Africa.

## Data Availability

All relevant data are within the manuscript and its supporting information files.

## Author Contributions

Conceived and designed the experiments: RAT, EP. Data Collection: RAT, AAY, EB. Performed the experiments: RAT, AAY, EB. Analyzed the data: RAT. Original Draft: RAT. Reviewed manuscript drafts: RAT, VS, LEA, EP.

## Acknowledgements

We thank the study participants, their parents or guardians, ART clinic staff, and the Ghana National AIDS Control Program for their cooperation and support. This study received support from the Fogarty International Center and the National Institute of Alcohol Abuse and Alcoholism (NIH D43 TW011526).

## Conflicts of Interest

The authors declare no conflicts of interest.

## Notes

### Competing Interest Statement

The authors have declared no competing interest.

### Funding Statement

Yes

### Author Declarations

approved by the Institutional Review Boards of the Ghana Health Service (Protocol # GHS-ERC: 025/09/21) and the Noguchi Memorial Institute for Medical Research, University of Ghana (Protocol # CPN 015/21-22).

